# The Association of Postoperative Anaemia with Outcomes in Cardiac Surgical Patients Eligible for Patient Blood Management: A Single Institution Retrospective Cohort Study

**DOI:** 10.1101/2022.08.21.22279032

**Authors:** Justyna Bartoszko, Michelle Li, Jeannie Callum, Sujung Yi, Maral Ouzounian, Stuart A. McCluskey, Sarah Miles, Yulia Lin, Keyvan Karkouti

**Affiliations:** Institute of Medical Science, University of Toronto, Toronto, Ontario, Canada; Department of Anesthesia and Pain Management, Sinai Health System, Women’s College Hospital, University Health Network, Toronto, Ontario, Canada; Department of Anesthesiology and Pain Medicine, University of Toronto, Toronto, Ontario, Canada; Peter Munk Cardiac Centre, University Health Network, Toronto, Ontario, Canada; University of Toronto Quality in Utilization, Education and Safety in Transfusion Research Program, Toronto, Ontario, Canada; Department of Pathology and Molecular Medicine, Kingston Health Sciences Centre and Queen’s University, Kingston, Ontario, Canada; Division of Cardiac Surgery, Department of Surgery, University of Toronto, Toronto, Ontario, Canada; Precision Diagnostics and Therapeutics Program, Sunnybrook Health Sciences Centre, Toronto, Ontario, Canada; Department of Laboratory Medicine and Pathobiology, University of Toronto, Toronto, Ontario, Canada; Institute for Health Policy, Management, and Evaluation, University of Toronto, Toronto, Ontario, Canada; Interdepartmental Division of Critical Care, Department of Medicine, University of Toronto, Toronto, Ontario, Canada

## Abstract

**Background:** Anaemia is prognostically important and affects 30-40% of cardiac surgical patients. The objective of this study was to examine the association of pre- and postoperative anaemia with outcomes in cardiac surgical patients.

**Methods:** This was a single-institution retrospective cohort study including cardiac surgical patients from October 26, 2020 to December 3, 2021. Patients were classified as preoperatively non-anaemic (hemoglobin ≥ 130 g/L), anaemic, or treated with IV Iron. The main predictors of interest were nadir haemoglobin on postoperative days 1-2 and preoperative anaemia and receipt of IV iron therapy. The primary outcome was number of red blood cell units (RBC) transfused on postoperative days 1-7. Secondary outcomes included acute kidney injury, hospital length of stay, and 30 day in-hospital mortality. Regression models, adjusted for demographics, comorbidities, and surgical characteristics, examined the association between predictors and outcomes.

**Results:** A total of 844 patients were included [528 (63%) non-anaemic, 276 (33%) anaemic, and 40 (5%) anaemic, treated with IV iron]. There was no difference between groups in RBC transfusion or mortality, however anaemic patients had a higher adjusted risk for acute kidney injury [aOR 2.69 (95% CI, 1.37 to 5.30), p=0.004] and longer hospital length of stay [aRR 1.38 (95% CI, 1.24 to 1.54), p<0.0001] compared to non-anaemic patients. Patients treated with IV iron did not have the same increased risk. A lower postoperative haemoglobin nadir was significantly associated with increased risk for all outcomes.

**Conclusions:** Postoperative anaemia confers additional risk regardless of preoperative anaemia status. Further research is needed to better clarify these associations.

## INTRODUCTION

Anaemia is a common and prognostically important comorbidity, affecting 30-40% of cardiac surgical patients.^1-3^ The presence of preoperative anaemia, defined as a haemoglobin level less than 120-130 g/L^4^, increases the risk-adjusted odds of major organ dysfunction, prolonged hospitalization, readmission, and death up to 4-fold compared to non-anaemic cardiac surgical patients.^3, 5^ Given the significant blood loss associated with open cardiac surgical procedures, anaemia becomes even more common following surgery, occurring in up to 90% of patients.^1, 6, 7^

The primary cause of preoperative anaemia in cardiac surgical patients is iron-deficiency, resulting in iron-restricted erythropoiesis.^2, 8^ Cardiac surgery exacerbates iron-deficiency for several weeks to months as it causes a systemic inflammatory response that enhances iron sequestration along with a state of acute-on-chronic anemia that increases erythropoiesis demand to a degree that can outstrip the available iron supply.^9, 10^

Given that iron-deficiency anaemia is highly prevalent and prognostically important in surgical patients, it has been flagged as a promising therapeutic target to improve surgical outcomes.^8^ Indeed, preoperative IV iron therapy to treat iron-deficiency anaemia has become the cornerstone of Patient Blood Management (PBM), a World Health Organization-supported initiative to ‘improve clinical outcomes by promoting best practices in transfusion medicine.’^11^ Oral iron supplements are often not effective in surgical patients because they are poorly tolerated^12^ and have low bioavailability.^13, 14^ IV iron formulations, on the other hand, have superior side-effect profiles and can safely deliver ≥1000 mg of elemental iron in a single infusion.^15, 16^ However, despite enthusiasm for the adoption of preoperative IV iron therapy in PBM programs, several systematic reviews across a range of surgeries, patient populations, and iron formulations have not consistently demonstrated that preoperative IV iron therapy improves surgical outcomes in those patients receiving it.^17-19^

There has also been growing interest in the association between postoperative anaemia and adverse events following cardiac surgery. Recently several studies have highlighted the important association of postoperative anaemia with outcomes, including acute kidney injury, major ischemic events, disability-free survival, and quality of recovery.^4, 20-23^ There is a need to further delineate and clarify these associations - particularly the impact that interventions such as preoperative IV iron therapy may have on postoperative anaemia.

We hypothesize that postoperative anaemia has a strong prognostic association with outcomes in cardiac surgical patients, which may be attenuated by access to preoperative IV iron. We conducted a single institution retrospective cohort study to assess the association of postoperative anaemia on rates of postoperative red blood cell transfusion, acute kidney injury, hospital length of stay, and 30 day in-hospital mortality. The modifying impact of preoperative IV iron was examined on postoperative haemoglobin levels and clinical outcomes.

## METHODS

This manuscript is reported in accordance with the STrengthening the Reporting of OBservational studies in Epidemiology (STROBE) statement.^24^ Institutional ethics approval to conduct the study was obtained from the University Health Network Research Ethics Board (Study Number 21-6193).

### Study Design

This retrospective cohort study was undertaken to examine differences in postoperative outcomes related to postoperative anaemia in patients undergoing elective or urgent cardiac surgery at a single institution, while assessing effect modification related to preoperative anaemia status and exposure to IV iron. We selected Hb <130 g/L as the cut-off for anaemia for both males and females as current guidelines recommending a lower cut-off of Hb <120 g/L for females may contribute to poorer outcomes in females.^1, 25-28^ Guidelines recommending a lower Hb cut-off in women (<120 g/L) were based on few studies, did not adequately account for different life stages in women (after menopause, mean hemoglobin levels increase to 135-140 g/L among women in their 50’s to 60’s), and did not exclude populations with iron-deficiency.^29^

### Patient Population and Data Sources

All patients undergoing elective cardiac surgery (defined as a stable clinical status for days or weeks prior to the operation) or urgent cardiac surgery (defined as worsening or sudden chest pain, heart failure, acute myocardial infarction, compelling anatomy, unstable angina or rest angina) at the University Health Network and enrolled in the Society for Thoracic Surgeons (STS) National Database for Cardiac Surgery at our institution between October 26, 2020 to December 3, 2021 were eligible for inclusion. Cases are assessed by a local PBM nurse at the time of referral to the pre-surgical assessment clinic according to the Ontario Nurse Transfusion Coordinators (ONTraC) algorithm (Supplemental Appendix 1). For anaemic patients undergoing elective or urgent surgery iron indices (serum ferritin, serum iron, transferrin saturation, and C-reactive protein) were ordered in addition to routine preoperative bloodwork. Patients with anaemia and iron deficiency are offered IV iron (300 mg iron sucrose for up to two doses, or ferric derisomaltose 500 mg if ≤ 60 kg or 1000 mg if > 60 kg for one dose) prior to surgery. PBM intervention-related data was obtained from local treatment records. Patient demographics, transfusion, and laboratory data were obtained from our institutional electronic health record. Patient comorbidity, procedure-related data, and outcomes were obtained from both our institutional electronic health record as well as our institution’s local STS Cardiac Surgery database.

### Predictors and Outcomes

The primary predictors of interest were nadir hemoglobin concentration on postoperative days 1 through 2 and preoperative anaemia status, including receipt of IV iron delivered as an intervention for the treatment of preoperative anaemia via the institutional PBM program. Preoperative anaemia as a predictor was defined using the most recent haemoglobin available prior to index surgery. The primary outcome of interest was the number of red blood cell (RBC) units transfused postoperatively from days 1 through 7. Secondary outcomes included the occurrence of Stage 1 or greater acute kidney injury (AKI) according to the Kidney Disease Improving Global Outcomes (KDIGO)^30^ grading system corresponding to a serum creatinine increase ≥1.5 times baseline, in-hospital 30 day mortality, and total hospital length of stay (with censoring of patients who had expired while in hospital).

### Statistical Analysis

Patients were divided into three groups, those who had no diagnosis of preoperative anaemia, those who received preoperative IV iron, and those with preoperative anaemia who did not receive IV iron. Continuous variables are expressed as medians and interquartile ranges. Categorical variables are reported as counts and percentages. Patient demographic, procedural, and laboratory characteristics were compared between the three pre-specified groups, which capture preoperative anaemia status and receipt of IV iron. The Kruskal-Wallis test was used for three-group comparisons of medians for continuous data, while the Wilcoxon Two-Sample test was used for comparisons of medians across two groups only. Analysis of Variance (ANOVA) was used for three-group comparisons of means, while the t-test was used for comparisons of means across two groups. The Chi-Square Test or Fisher’s Exact test (for cell counts < 5) were used to compare count data. The association between postoperative nadir haemoglobin level and outcomes was visually examined using scatterplots.

Postoperative haemoglobin was treated as a continuous variable to obtain an odds or risk ratio for outcomes per 10 g/L decrease. Unadjusted and adjusted logistic regression models were used to assess the association between predictors of interest (postoperative day 1-2 nadir haemoglobin and preoperative anaemia status) and binary outcomes. Unadjusted and adjusted negative binomial regression models were used to assess the association between predictors of interest and dispersed count data. Models were examined to ensure model fit and assumptions were met. For each logistic regression model, we used global measures of model fit based on information criterion statistics. Model discrimination was assessed using the C-statistic. Model calibration was assessed through the use of the Hosmer-Lemeshow test by comparing the observed events within prespecified risk groupings (deciles of risk), with higher P values indicating better calibration. For negative binomial models, the Akaike Information Criterion was used for model selection and calibration.^31^

According to a recent conceptual model proposed for the association of postoperative anaemia with outcomes, confounders were specified *a priori* based on their clinical relevance and known impact on postoperative events in the literature.^4^ Models were adjusted for procedure complexity (defined as complex if more than just CABG alone, single valve surgery alone, or ASD repair alone), age, sex, preoperative dialysis, preoperative serum creatinine level, the presence of chronic lung disease, liver disease, cerebrovascular disease, prior myocardial infarction, heart failure, urgent or elective status, and whether the index surgery was a redo procedure. All clinically important variables showed no multi-collinearity (correlation coefficients < 0.50) and were entered into the multivariable regression models.

Special data handling methods were not employed for missing data, as generally there was minimal missing data in the dataset (<10%). SAS Studio (SAS Institute, Inc, Cary, NC) was used for all statistical analyses. All reported P-values were 2 sided, and values of P<0.05 were considered to be statistically significant.

## RESULTS

### Patient Characteristics

A total of 844 patients were eligible for inclusion in our study, with 528 (63%) non-anaemic properatively, 277 (33%) anaemic with no IV iron treatment preoperatively, and 39 (5%) having received IV iron prior to surgery. Preoperative haematocrit and haemoglobin levels were similar between patients in the anaemic group with no treatment and the group that received preoperative IV iron (Table 1), and both these groups had a significantly lower haemoglobin than the non-anaemic group. Patients in the anaemic and IV iron groups were older with a higher proportion of females and patients with dialysis dependence. A greater proportion of patients with untreated anaemia underwent urgent compared to elective surgery while the reverse was true for non-anemic and untreated anemic patients. In the non-anaemic group, the median postoperative nadir haemoglobin was 94 g/L (IQR, 82-106), in the untreated anaemic group it was 81 g/L (IQR, 75-90), and in the IV iron group it was 85 g/L (IQR, 78-91), (3 group Kruskal-Wallis p<0.0001; Anaemic vs. Treated Wilcoxon p=0.27). The median haemoglobin drop from the last preoperative haemoglobin to the nadir haemoglobin on postoperative days 1-2 was 51 g/L (IQR, 41-63) in the non-anaemic group, 31 g/L (IQR 22-41) in the anaemic group, and 33 g/L (IQR, 20-43) in the IV iron group, (3 group Kruskal-Wallis p<0.0001; Anaemic vs. Treated Wilcoxon p=0.44). Anaemic patients received a median of 2 (IQR, 0-3) RBC units on the day of surgery, while patients treated with IV iron received a median of 2 (IQR, 0-3), (Wilcoxon p=0.64). Overall, patients in the IV Iron group tended to cluster with the untreated anaemic patients in terms of their preoperative haemoglobins, their transfusion needs on the day of surgery, and their postoperative nadir haemoglobins (Figure 1). There was a moderate correlation between preoperative and postoperative nadir haemoglobin which was maintained across all patient groups (Figure 1, Pearson rho=0.49, p<0.0001).

**Table 1.** Patient Characteristics.

|  | **Non-Anemic**<br>**(n=528)** | **Anemia, no IV Iron**<br>**(n=277)** | **Preoperative**<br>**IV Iron**<br>**(n=39)** |  |
| --- | --- | --- | --- | --- |
|  | **Demographic Characteristics and Preoperative Laboratory values** |  |  |  |
| Last Preoperative Hemoglobin Level (g/L), median (IQR) | 146 (139, 154) | 117 (103, 125) | 120 (110, 127) | <0.0001 |
| Preoperative Hematocrit, median (IQR) | 44 (42, 46) | 36 (32, 38) | 37 (35, 40) | <0.0001 |
| Age (years), median (IQR) | 61 (51, 68) | 66 (57, 73) | 68 (52, 72) | <0.0001 |
| Last Preoperative Serum Creatinine Level (μmol/L), median (IQR) | 85 (74, 96) | 87 (71, 120) | 80 (70, 97) | 0.06 |
| Last preoperative INR, median (IQR) | 1.1 (1.0, 1.1) | 1.1 (1.1, 1.2) | 1.1 (1.0, 1.2) | <0.0001 |
| Height (cm), median (IQR) | 173 (167, 180) | 168 (160, 175) | 165 (162, 171) | <0.0001 |
| Weight (kg), median (IQR) | 83 (71, 95) | 78 (64, 88) | 76 (61, 86) | <0.0001 |
| Sex (Female), n (%) | 97 (18%) | 116 (44%) | 22 (61%) | <0.0001 |
|  | **Comorbidities** |  |  |  |
| Diabetes, n (%) | 106 (20%) | 93 (34%) | 6 (15%) | <0.0001 |
| Dialysis Dependence, n (%) | 9 (2%) | 26 (9%) | 1 (3%) | <0.0001 |
| Hypertension, no (%) | 309 (59%) | 177 (64%) | 16 (41%) | 0.008 |
| Tobacco Use, Current, n (%) | 70 (13%) | 37 (13%) | 3 (8%) | 0.55 |
| Chronic Lung Disease, n (%) | 22 (4%) | 17 (6%) | 1 (3%) | 0.47 |
| Alcohol Use, n (%) | 248 (47%) | 111 (40%) | 9 (23%) | 0.002 |
| Liver Disease, n (%) | 8 (2%) | 3 (1%) | 1 (3%) | 0.57 |
| Immunocompromised, n (%) | 9 (2%) | 11 (4%) | 3 (8%) | 0.03 |
| Cancer in Last 5 Years, n (%) | 13 (2%) | 14 (5%) | 1 (3%) | 0.15 |
| Peripheral Arterial Disease, n (%) | 20 (4%) | 14 (5%) | 0 (0%) | 0.32 |
| Cerebrovascular Disease, n (%) | 25 (5%) | 25 (9%) | 3 (8%) | 0.06 |
| Prior Cerebrovascular Accident, n (%) | 9 (2%) | 8 (3%) | 3 (8%) | 0.11 |
| Previous Cardiac Intervention, n (%) | 123 (23%) | 70 (25%) | 9 (23%) | 0.87 |
| Previous Myocardial Infarction, n (%) | 51 (10%) | 34 (12%) | 4 (10%) | 0.59 |
| Myocardial Infarction Timing, n (%) |  |  |  | 0.15 |
| <21 days | 15 (3%) | 15 (5%) | 0 (0%) |  |
| ≥ 21 days | 36 (7%) | 19 (7%) | 4 (10%) |  |
| **Primary Coronary Symptoms, n (%)** |  |  |  |  |
| *None* | 150 (28%) | 82 (29%) | 15 (38%) | <0.0001 |
| *Stable Angina* | 177 (34%) | 58 (21%) | 20 (51%) |  |
| *Unstable Angina* | 83 (16%) | 77 (28%) | 2 (5%) |  |
| *NSTEMI* | 19 (4%) | 29 (11%) | 0 (0%) |  |
| *STEMI* | 12 (2%) | 8 (3%) | 0 (0%) |  |
| *Angina Equivalent* | 29 (5%) | 6 (2%) | 1 (3%) |  |
| *Other* | 9 (2%) | 3 (1%) | 0 (0%) |  |
| Heart Failure, n (%) | 41 (8%) | 49 (18%) | 12 (30%) | <0.0001 |
| Cardiogenic Shock at Time of Procedure | 1 (<1%) | 6 (2%) | 0 (0%) | 0.02 |
| Reoperation, n (%) | 59 (11%) | 28 (10%) | 9 (23%) | 0.07 |
| **Surgical Urgency, n (%)** |  |  |  |  |
| *Elective* | 391 (74%) | 122 (44%) | 34 (87%) | <0.0001 |
| *Urgent* | 137 (26%) | 155 (56%) | 5 (13%) |  |

*Kruskal-Wallis Test for continuous variables and Chi-Square test or Fisher's Exact test for cell counts <5*

**Figure 1.**
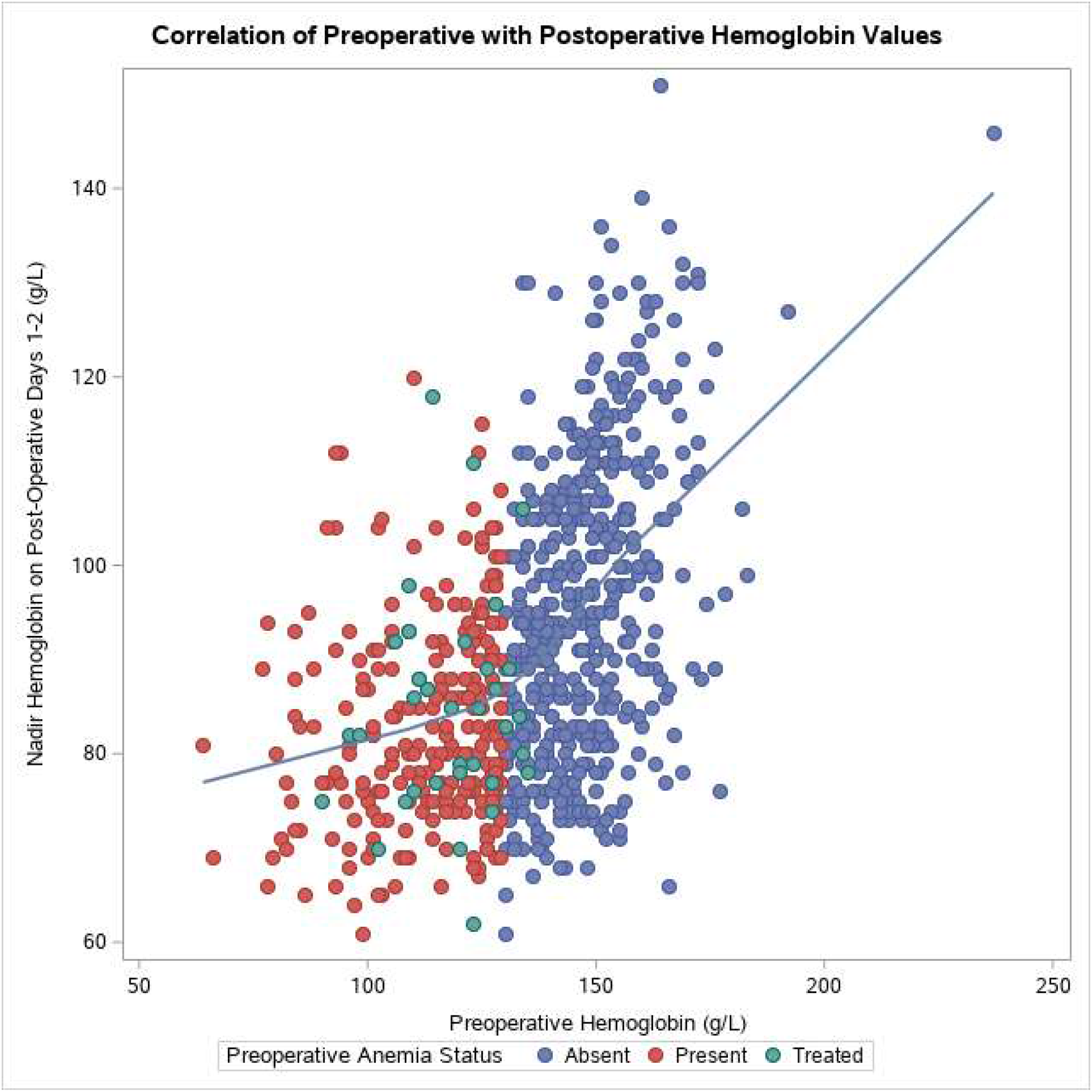
Correlation of Last Preoperative Serum Hemoglobin Level with Postoperative Day 1 to 2 Nadir Hemoglobin Level, by Preoperative Anemia and Treatment Group, Spearman Correlation Coefficient=0.49, p<0.001.

### Primary Outcome

A total of 708 units of red cells were transfused from postoperative days 1 through 7. Patients in the non-anaemic group received a mean of 0.59 (SD 1.39) RBC over days 1 through 7, patients in the anaemic group received a mean of 1.45 (SD 1.84), and patients in the IV Iron group received 0.81 (SD 0.95) (ANOVA, p<0.0001). The median number of RBC units transfused from days 1 through 7 was 0 (IQR, 0-1) in the non-anaemic group, 1 (IQR, 0-2) in the anaemic group, and 0.5 (IQR, 0-1.5) in the IV iron group (Kruskal-Wallis, p<0.0001). There was a trend towards significance between the anaemic and IV iron groups, with the IV iron group requiring less RBC transfusion during this time (Wilcoxon Two-Sample Test, p=0.06). Adjusted negative binomial regression found no difference between the non-anaemic, anaemic, and IV iron groups for the number of RBC units transfused during this time (Table 2i).

**Table 2.** Model Results for Primary and Secondary Outcomes.

**i.) Negative Binomial Regression Results for Outcome: Number of Red Blood Cell Units Transfused from Postoperative Day 1 through 7**
| **Outcome: Number of Red Blood Cell Units Transfused from Postoperative Day 1 through 7** |  |  |  |
| --- | --- | --- | --- |
| **Number of Red Cells (%)** | **Group** | **Unadjusted Relative Risk with 95% Confidence Interval** | **Adjusted Relative Risk with 95% Confidence Interval** |
| **Predictor: Preoperative Anemia and Treatment Status** |  |  |  |
| 300 (42%) | Non-Anemic | Reference |  |
| 379 (53%) | Anemia, no IV Iron | **2.44 (1.94 to 3.10)** | 1.18 (0.91 to 1.54) |
| 29 (4%) | Preoperative IV Iron | 1.36 (0.78 to 2.38) | 0.77 (0.45 to 1.30) |
| **Predictor: Hemoglobin Nadir on Postoperative Day 1 through 2** |  |  |  |
| 708 (100%) | Hemoglobin Level per 10 g/L decrease | **1.28 (1.21 to 1.36)** | **2.54 (2.25 to 2.88)** |

| **Outcome: KDIGO Stage 1 or Greater AKI** |  |  |  |
| --- | --- | --- | --- |
| **Proportion of Patients with AKI (%)** | **Group** | **Unadjusted Relative Risk with 95% Confidence Interval** | **Adjusted Relative Risk with 95% Confidence Interval** |
| **Predictor: Preoperative Anemia and Treatment Status** |  |  |  |
| 55 of 528 (10%) | Non-Anemic | Reference |  |
| 45 of 276 (16%) | Anemia, no IV Iron | **1.67 (1.09 to 2.55)** | **2.67 (1.36 to 5.26)** |
| 6 of 40 (15%) | Preoperative IV Iron | 1.56 (0.63 to 3.90) | 0.96 (0.24 to 3.85) |
| **Predictor: Hemoglobin Nadir on Postoperative Day 1 through 2** |  |  |  |
| 106 (100%) | Hemoglobin Level per 10 g/L decrease | **1.51 (1.27 to 1.80)** | **1.31 (1.05 to 1.66)** |

**iii.) Logistic Regression Results for Outcome: 30 Day In-Hospital Mortality**
| **Outcome: 30 Day In-Hospital Mortality** |  |  |  |
| --- | --- | --- | --- |
| **Number (%)** | **Group** | **Unadjusted Relative Risk with 95% Confidence Interval** | **Adjusted Relative Risk with 95% Confidence Interval** |

| Predictor: Preoperative Anemia and Treatment Status |  |  |  |
| --- | --- | --- | --- |
| 10 of 528 (2%) | Non-Anemic | Reference |  |
| 17 of 276 (6%) | Anemia, no IV Iron | **3.39 (1.53 to 7.50)** | 1.73 (0.44 to 6.74) |
| 1 of 40 (3%) | Preoperative IV Iron | 1.36 (0.17 to 10.93) | 1.20 (0.11 to 12.54) |
| Predictor: Hemoglobin Nadir on Postoperative Day 1 through 2 |  |  |  |
| 28 (100%) | Hemoglobin Level per 10 g/L increase | **1.77 (1.22 to 2.56)** | **1.69 (1.00 to 2.85)** |

**iv.) Unadjusted Logistic Regression Results for Outcome: Hospital Length of Stay**
| Outcome: Hospital Length of Stay |  |  |  |
| --- | --- | --- | --- |
| Mean Length of Stay in Days (SD) | Group | Unadjusted Relative Risk with 95% Confidence Interval | Adjusted Relative Risk with 95% Confidence Interval |
| Predictor: Preoperative Anemia and Treatment Status |  |  |  |
| 8.4 (7.8) | Non-Anemic | Reference |  |
| 14.3 (13.8) | Anemia, no IV Iron | **1.69 (1.54 to 1.87)** | **1.37 (1.23 to 1.54)** |
| 11.3 (9.5) | Preoperative IV Iron | **1.36 (1.09 to 1.70)** | 1.19 (0.97 to 1.46) |
| Predictor: Hemoglobin Nadir on Postoperative Day 1 through 2 |  |  |  |
| 10.4 (10.5) | Hemoglobin Level per 10 g/L increase | **1.13 (1.09 to 1.16)** | **1.03 (1.00 to 1.07)** |

*Models were adjusted for procedure complexity (defined as complex if more than just CABG or valve surgery alone), age, sex, pre-operative dialysis, pre-operative serum creatinine, pre-operative anaemia and IV iron status, the presence of chronic lung disease, liver disease, cerebrovascular disease, prior myocardial infarction, heart failure, urgent or elective status, and whether the index surgery was a redo procedure.*

There was a moderate to strong negative correlation between postoperative nadir haemoglobin level and units of RBC transfused from postoperative days 1 through 7 (Figure 2a, Spearman Rho=-0.73, p<0.0001). Postoperative nadir haemoglobin was strongly associated with RBC transfusion [Table 2i, aOR 2.54, 95% CI (2.25 to 2.88)].

**Figure 2.**
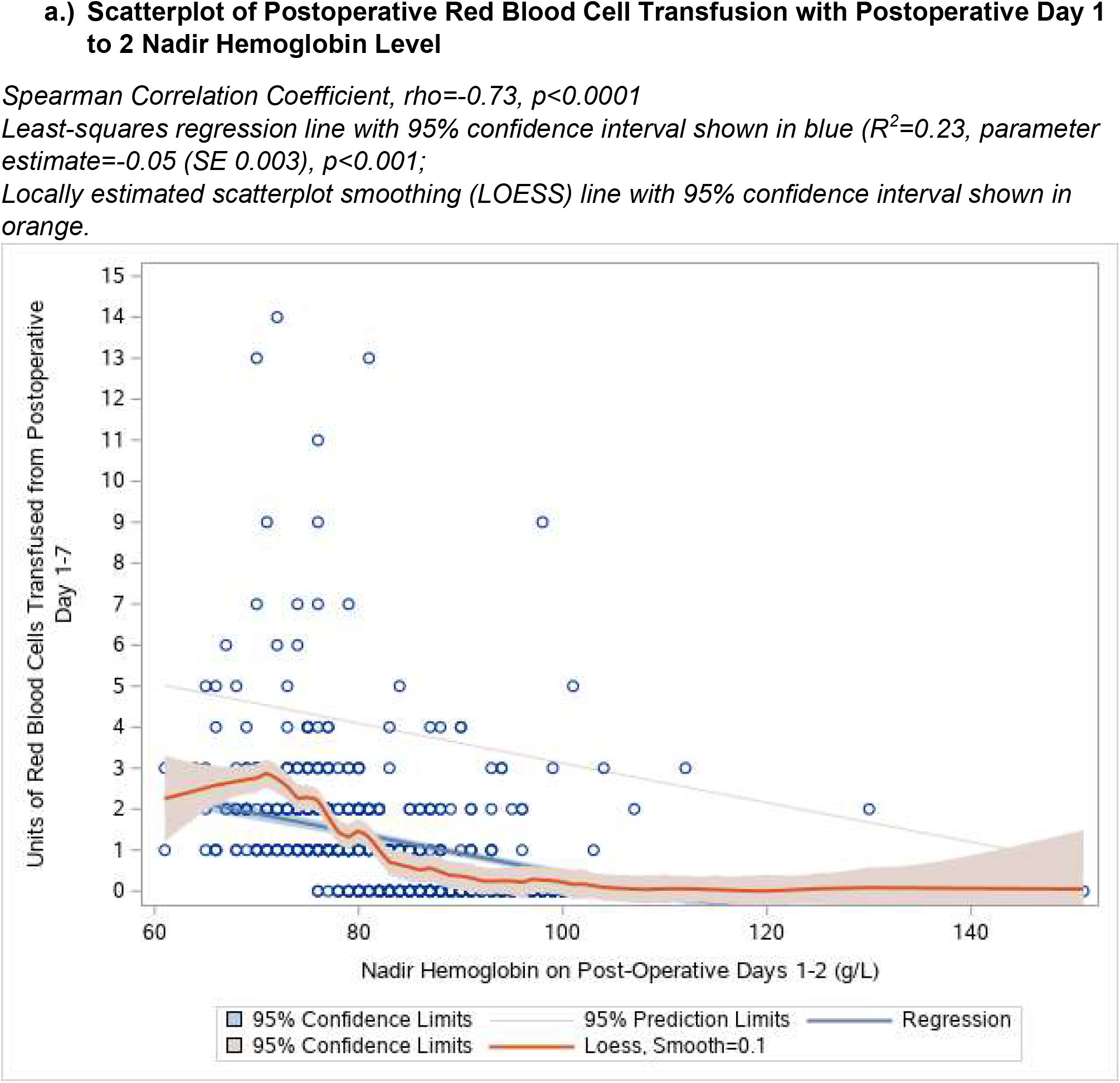
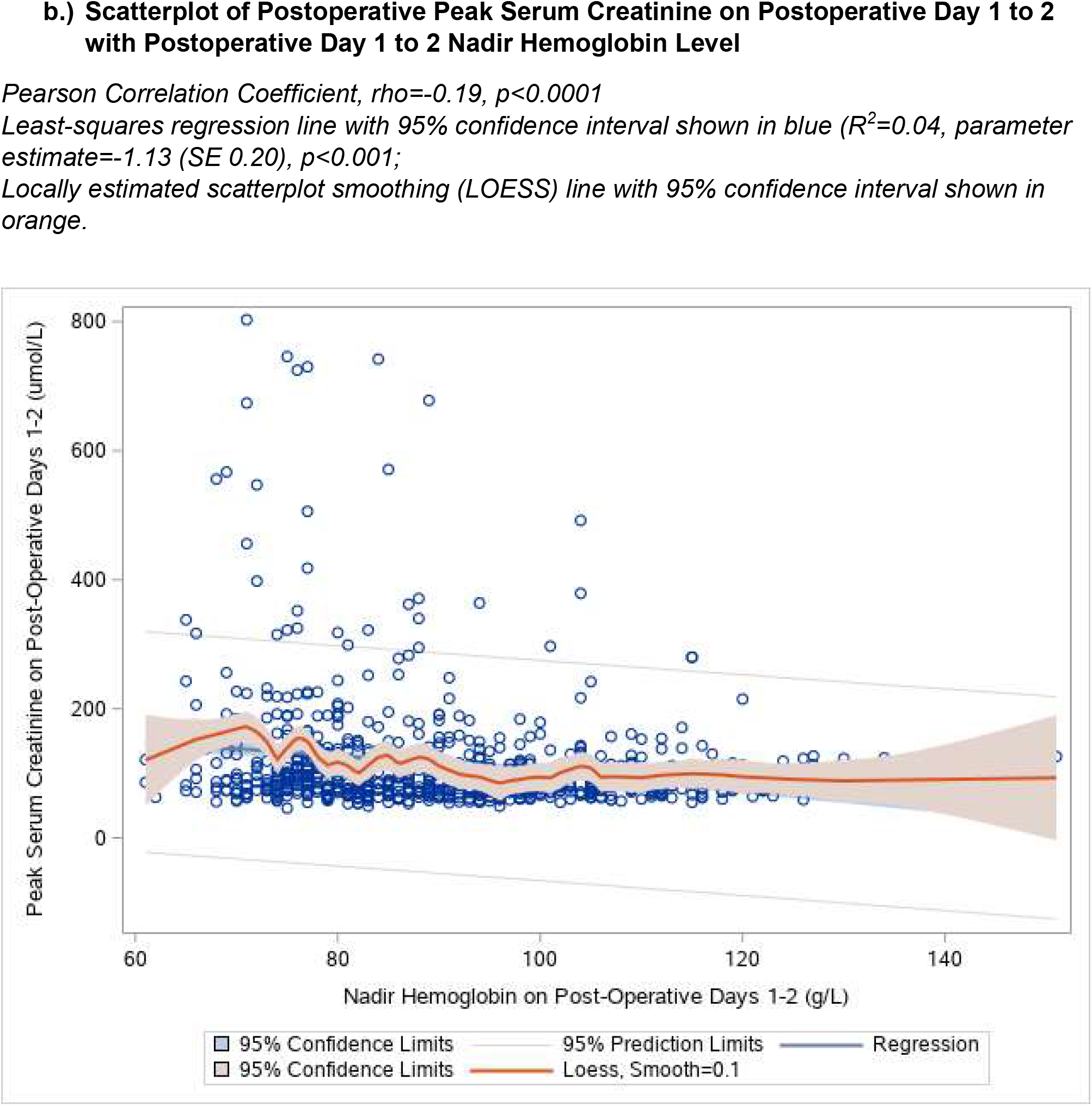
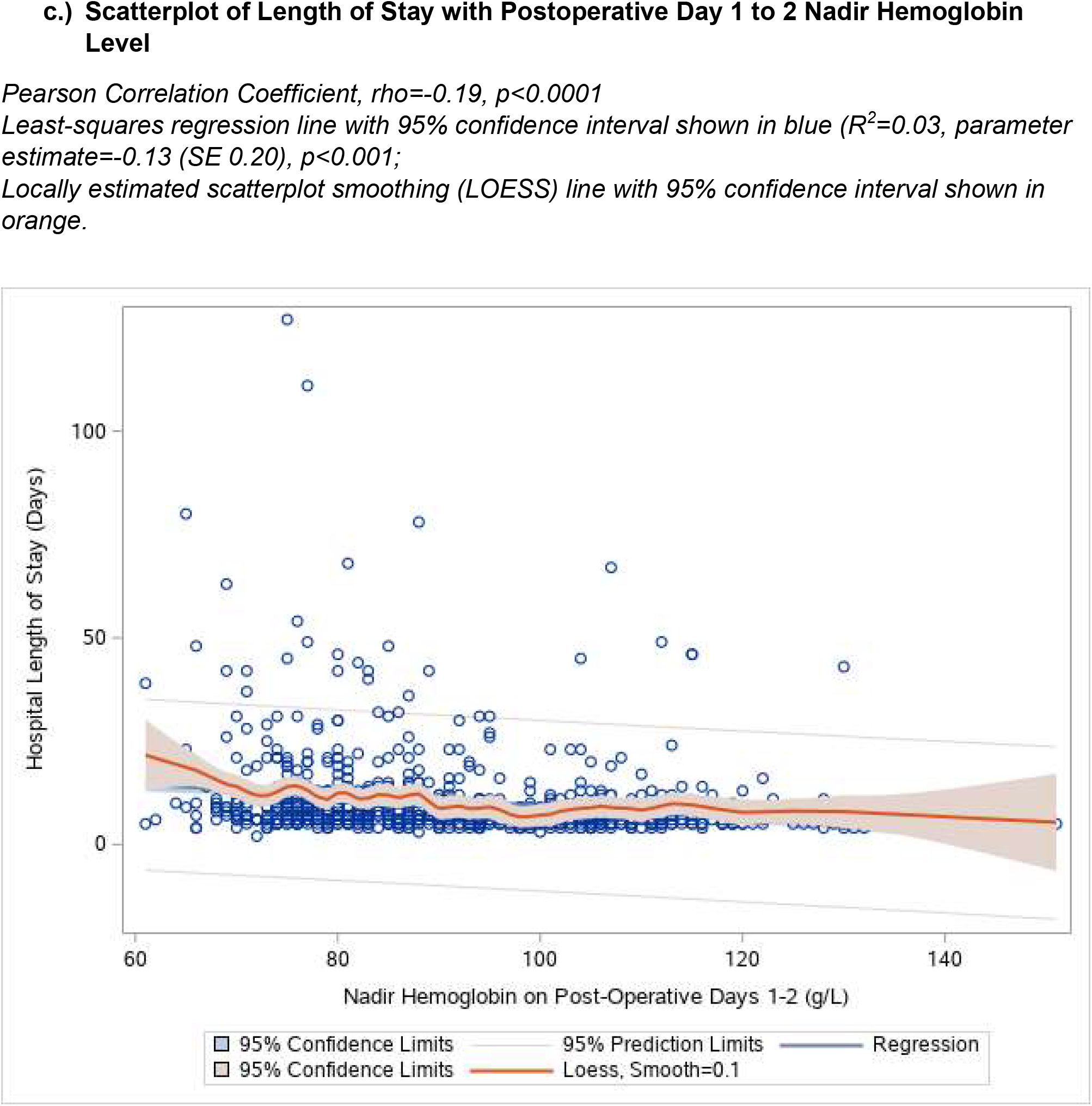
Associations Between Postoperative Day 1 to 2 Nadir Hemoglobin with Postoperative Red Blood Cell Transfusion, Peak Postoperative Serum Creatinine, and Length of Stay.

### Secondary Outcomes

#### Acute Kidney Injury

Stage 1 AKI according to the KDIGO classification^30^ occurred in 106 (13%) patients (Table 2ii). Across all patients groups, peak serum creatinine on postoperative days 1 through 2 was weakly associated with nadir haemoglobin levels (Figure 2b, Pearson rho=-0.19, p<0.0001). Compared to the non-anaemic group, anaemic patients had a significantly higher risk of Stage 1 AKI [aOR 2.67, 95% CI (1.36 to 5.26)], while the IV iron group was not significantly different from the non-anaemic group [aOR 0.96, 95% CI (0.24 to 3.85)] (Table 3ii). Lower postoperative haemoglobin level remained significantly associated with increased Stage 1 AKI [aOR per 10 g/L haemoglobin decrease 1.31, 95% CI (1.05 to 1.66)] (Table 3ii).

#### 30 Day In-Hospital Mortality

A total of 28 (3%) patients experienced in-hospital mortality within 30 days of surgery. Both anaemic patients and patients treated with IV iron were not found to have a significantly higher risk of mortality than non-anaemic patients in adjusted or unadjusted models (Tables 2iii). While nadir postoperative haemoglobin remained associated with mortality, this association was weakened in the adjusted models [aOR per 10 g/L haemoglobin decrease 1.69, 95% CI (1.00 to 2.85)] (Table 2iii).

#### Hospital Length of Stay

The overall mean length of stay was 10.4 days (SD 10.5). Postoperative nadir haemoglobin was weakly correlated with hospital length of stay when examined across all patients (Figure 2d, Pearson Correlation Coefficient, rho=-0.19, p<0.0001). In unadjusted negative binomial models, both the untreated anaemic and IV iron groups had a longer length of stay than the non-anaemic groups (Table 2iv). After adjustment, a strong association persisted only for the untreated anaemic group [aRR 1.37, 95% CI (1.23 to 1.54)]. Compared to the non-anaemic group, patients who received IV iron did not have a significantly different length of stay after adjustment for risk factors [aRR 1.19, 95% CI (0.97 to 1.46)] (Table 2iv). Nadir postoperative haemoglobin levels remained associated with hospital length of stay in both unadjusted and adjusted models [aRR per 10 g/L haemoglobin decrease 1.03, 95% CI (1.00 to 1.07)] (Table 2iv).

## DISCUSSION

This single-institution retrospective cohort study demonstrates the association of postoperative haemoglobin level with adverse outcomes in patients undergoing elective and urgent cardiac surgery, while reaffirming the additional adverse prognostic impact of preoperative anaemia which remains untreated. This study suggests that patients who received preoperative IV iron as a treatment for anaemia may benefit from a reduction in the risk of acute kidney injury and a reduction in the overall length of stay. However, all patients – whether anaemic and untreated, treated with IV iron, or non-anaemic – were susceptible to an additional risk of adverse outcomes related to the degree of postoperative anaemia.

As with other acute illnesses, the postoperative period is a period of heightened vulnerability for patients,^32^ particularly those with anaemia. As confirmed in this study, cardiac surgery exacerbates anaemia quite significantly postoperatively,^9, 10, 33-35^ causing haemoglobin levels to drop by an average of 30-40% and remain below 100 g/L for several weeks to months.^36, 37^ During this phase of postoperative recovery, patient vulnerability to adverse outcomes is heightened due to impairments in end-organ function, cardiorespiratory fitness, and physical performance.^36, 38^ Among patients within one year of hospitalization for surgery and critical illness, each monthly 10 g/L recovery in haemoglobin is associated with a lower instantaneous hazard for readmission (adjusted hazard ratio 0.87; 95% CI 0.84-0.90; p < 0.001) and mortality (adjusted hazard ratio 0.82; 95% CI 0.75-0.89; p < 0.001).^39^ Concurrently, and as this study demonstrates, this risk appears heightened in patients with preoperative anaemia which remains untreated.

While patients would ideally be treated for iron deficiency anaemia before surgery, prior studies have demonstrated the challenges of being able to do this effectively with short surgical lead times.^40^ In our sample, the median time patients received IV prior to surgery was only 5.5 days, IQR (2.0-10.5). An important finding of this study was the low rate of patients receiving preoperative IV iron (only 7% of all anaemic patients in this population). This is likely to be related to a short lead time prior to surgery and logistical constraints with arranging outpatient IV iron therapy (e.g., shortage of short-notice outpatient IV infusion appointments). Therefore, the immediate postoperative period provides a valuable window to treat iron-deficiency anaemia to improve clinical outcomes. Furthermore, given the vulnerability of patients with chronic anaemia to the risks of postoperative anaemia, and the known worsening and persistence of anaemia after hospitalization and surgery, it is likely that interventions aimed at managing anaemia need to be in effect for at least several weeks after surgery to improve clinical outcomes and reduce readmissions.

Limitations of this study relate to the retrospective study design. Despite adjustment for important confounders, anaemic patients tend to be sicker with a greater burden of comorbidity and the possibility of residual confounding exists. As our predictor of interest was the nadir hemoglobin on postoperative days 1 to 2, this may have been affected by intraoperative red blood cell transfusion prior to measurement which in turn may also impact outcomes. Additionally, our results are limited by the small number of patients in our cohort who received IV iron, decreasing our ability to detect small differences between groups. Despite these recognized limitations however, it is clear that patients treated with IV iron remained clustered with their untreated anaemic counterparts with respect to their pre- and postoperative haemoglobin levels, and that postoperative anaemia poses increased risk to all patients – including those who were not anaemic preoperatively.

Further research is required to better delineate the associations between postoperative anaemia and outcomes, including treatment regimens to address this widespread problem for which effective interventions (i.e. IV iron) exist. Future studies examining the impact of preoperative as well as postoperative IV iron with the aim of improving patient outcomes are warranted.

## Data Availability

All data produced in the present work are contained in the manuscript.

## REFERENCES

1 Muñoz M, Acheson AG, Auerbach M, et al. International consensus statement on the peri-operative management of anaemia and iron deficiency. Anaesthesia 2017; 72: 233–47

2 Hung M, Ortmann E, Besser M, et al. A prospective observational cohort study to identify the causes of anaemia and association with outcome in cardiac surgical patients. Heart 2015; 101: 107–12

3 Karkouti K, Wijeysundera DN, Beattie WS. Risk associated with preoperative anemia in cardiac surgery: a multicenter cohort study. Circulation 2008; 117: 478–84

4 Myles PS, Richards T, Klein A, et al. Postoperative anaemia and patient-centred outcomes after major abdominal surgery: a retrospective cohort study. Br J Anaesth 2022

5 Iribarne A, Chang H, Alexander JH, et al. Readmissions after cardiac surgery: experience of the National Institutes of Health/Canadian Institutes of Health research cardiothoracic surgical trials network. Ann Thorac Surg 2014; 98: 1274–80

6 Gómez-Ramírez S, Bisbe E, Shander A, Spahn DR, Muñoz M. Management of Perioperative Iron Deficiency Anemia. Acta Haematologica 2019; 142: 21–9

7 Rössler J, Schoenrath F, Seifert B, et al. Iron deficiency is associated with higher mortality in patients undergoing cardiac surgery: a prospective study. Br J Anaesth 2020; 124: 25–34

8 Muñoz M, Laso-Morales MJ, Gómez-Ramírez S, Cadellas M, Núñez-Matas MJ, García-Erce Ja. Pre-operative haemoglobin levels and iron status in a large multicentre cohort of patients undergoing major elective surgery. Anaesthesia 2017; 72: 826–34

9 Karkouti K, Yip P, Chan C, Chawla L, Rao V. Pre-operative anaemia, intra-operative hepcidin concentration and acute kidney injury after cardiac surgery: a retrospective observational study. Anaesthesia 2018; 73: 1097–102

10 Laffey JG, Boylan JF, Cheng DC. The systemic inflammatory response to cardiac surgery: implications for the anesthesiologist. Anesthesiology 2002; 97: 215–52

11 Spahn DR, Muñoz M, Klein AA, Levy JH, Zacharowski K. Patient Blood Management: Effectiveness and Future Potential. Anesthesiology 2020; 133: 212–22

12 Short MW, Domagalski JE. Iron deficiency anemia: evaluation and management. Am Fam Physician 2013; 87: 98–104

13 Stoffel NU, von Siebenthal HK, Moretti D, Zimmermann MB. Oral iron supplementation in iron-deficient women: How much and how often? Mol Aspects Med 2020; 75: 100865

14 Stoffel NU, Cercamondi CI, Brittenham G, et al. Iron absorption from oral iron supplements given on consecutive versus alternate days and as single morning doses versus twice-daily split dosing in iron-depleted women: two open-label, randomised controlled trials. Lancet Haematol 2017; 4: e524–e33

15 Wang C, Graham DJ, Kane RC, et al. Comparative Risk of Anaphylactic Reactions Associated With Intravenous Iron Products. JAMA 2015; 314: 2062–8

16 O’Lone EL, Hodson EM, Nistor I, Bolignano D, Webster AC, Craig JC. Parenteral versus oral iron therapy for adults and children with chronic kidney disease. Cochrane Database Syst Rev 2019; 2: Cd007857

17 Moon T, Smith A, Pak T, et al. Preoperative Anemia Treatment with Intravenous Iron Therapy in Patients Undergoing Abdominal Surgery: A Systematic Review. Advances in Therapy 2021; 38: 1447–69

18 Alshantti AH, Ahmed Z, Robertson S, Aboumarzouk OM, Alshantti A. Intravenous Iron versus Oral Iron in Anemia Management for Perioperative Patients: A Systemic Review and Meta-Analysis. Journal of Applied Hematology 2020; 11: 184–90

19 Tankard KA, Park B, Brovman EY, Bader AM, Urman RD. The Impact of Preoperative Intravenous Iron Therapy on Perioperative Outcomes in Cardiac Surgery: A Systematic Review. J Hematol 2020; 9: 97–108

20 Smoor RM, Rettig TCD, Vernooij LM, et al. Postoperative anaemia and disability-free survival in older cardiac surgery patients. British Journal of Anaesthesia 2022; 129: e27–e9

21 Cui R, Li F, Shao J, et al. Postoperative anemia is a risk factor for acute kidney injury after open aorta and vena cava surgeries. PLoS One 2020; 15: e0240243

22 Kougias P, Sharath S, Barshes NR, Chen M, Mills JL, Sr. Effect of postoperative anemia and baseline cardiac risk on serious adverse outcomes after major vascular interventions. Journal of Vascular Surgery 2017; 66: 1836–43

23 Zhou L, Liu X, Yan M, et al. Postoperative Nadir Hemoglobin and Adverse Outcomes in Patients Undergoing On-Pump Cardiac Operation. The Annals of Thoracic Surgery 2021; 112: 708–16

24 von Elm E, Altman DG, Egger M, et al. The Strengthening the Reporting of Observational Studies in Epidemiology (STROBE) statement: guidelines for reporting observational studies. Lancet 2007; 370: 1453–7

25 Dugan C, MacLean B, Cabolis K, et al. The misogyny of iron deficiency. Anaesthesia 2021; 76 Suppl 4: 56–62

26 Weyand AC, McGann PT, Sholzberg M. Sex specific definitions of anaemia contribute to health inequity and sociomedical injustice. Lancet Haematol 2022; 9: e6–e8

27 Blaudszun G, Munting KE, Butchart A, Gerrard C, Klein AA. The association between borderline pre-operative anaemia in women and outcomes after cardiac surgery: a cohort study. Anaesthesia 2018; 73: 572–8

28 Lin Y. Preoperative anemia-screening clinics. Hematology 2019; 2019: 570–6

29 Jorgensen JM, Crespo-Bellido M, Dewey KG. Variation in hemoglobin across the life cycle and between males and females. Annals of the New York Academy of Sciences 2019; 1450: 105–25

30 Levey AS, Eckardt KU, Dorman NM, et al. Nomenclature for kidney function and disease: report of a Kidney Disease: Improving Global Outcomes (KDIGO) Consensus Conference. Kidney Int 2020; 97: 1117–29

31 Portet S. A primer on model selection using the Akaike Information Criterion. Infect Dis Model 2020; 5: 111–28

32 Shawon MSR, Odutola M, Falster MO, Jorm LR. Patient and hospital factors associated with 30-day readmissions after coronary artery bypass graft (CABG) surgery: a systematic review and meta-analysis. Journal of Cardiothoracic Surgery 2021; 16

33 Gómez-Ramirez S, Jericó C, Muñoz M. Perioperative anemia: Prevalence, consequences and pathophysiology. Transfus Apher Sci 2019; 58: 369–74

34 Nairz M, Haschka D, Demetz E, Weiss G. Iron at the interface of immunity and infection. Front Pharmacol 2014; 5: 152

35 van Iperen CE, Kraaijenhagen RJ, Biesma DH, Beguin Y, Marx JJ, van de Wiel A. Iron metabolism and erythropoiesis after surgery. Br J Surg 1998; 85: 41–5

36 Zhou L, Liu X, Yan M, et al. Postoperative Nadir Hemoglobin and Adverse Outcomes in Patients Undergoing On-Pump Cardiac Operation. The Annals of thoracic surgery 2021; 112: 708–16

37 Karkouti K, Wijeysundera DN, Yau TM, McCluskey SA, Van Rensburg A, Beattie WS. The influence of baseline hemoglobin concentration on tolerance of anemia in cardiac surgery. Transfusion 2008; 48: 666–72

38 Kalra SK, Thilagar B, Khambaty M, Manjarrez E. Post-operative Anemia After Major Surgery: a Brief Review. Current Emergency and Hospital Medicine Reports 2021; 9: 89–95

39 Warner MA, Hanson AC, Schulte PJ, et al. Early Post-Hospitalization Hemoglobin Recovery and Clinical Outcomes in Survivors of Critical Illness: A Population-Based Cohort Study. J Intensive Care Med 2022: 8850666211069098

40 Richards T, Baikady RR, Clevenger B, et al. Preoperative intravenous iron to treat anaemia before major abdominal surgery (PREVENTT): a randomised, double-blind, controlled trial. The Lancet 2020; 396: 1353–61

